# Children and adults with mild COVID-19 symptoms develop memory T cell immunity to SARS-CoV-2

**DOI:** 10.1101/2021.09.10.21263333

**Authors:** Patricia Kaaijk, Verónica Olivo Pimentel, Maarten E. Emmelot, Martien Poelen, Alper Cevirgel, Rutger M. Schepp, Gerco den Hartog, Daphne F.M. Reukers, Lisa Beckers, Josine van Beek, Cécile A.C.M. van Els, Adam Meijer, Nynke Y. Rots, Jelle de Wit

## Abstract

**Background:** Severe acute respiratory syndrome coronavirus-2 (SARS-CoV-2) has led to considerable morbidity/mortality worldwide, but most infections, especially among children, have a mild course. However, it remains largely unknown whether infected children develop cellular immune memory.

**Methods:** To determine whether a memory T cell response is being developed as an indicator for long-term immune protection, we performed a longitudinal assessment of the SARS-CoV-2-specific T cell response by IFN-γ ELISPOT and activation marker expression analyses of peripheral blood samples from children and adults with mild-to-moderate COVID-19.

**Results:** Upon stimulation of PBMCs with heat-inactivated SARS-CoV-2 or overlapping peptides of spike (S-SARS-CoV-2) and nucleocapsid proteins, we found S-SARS-CoV-2-specific IFN-γ T cell responses in most infected children (83%) and all adults (100%) that were absent in unexposed controls. Frequencies of SARS-CoV-2-specific T cells were higher in infected adults, especially in those with moderate symptoms, compared to infected children. The S-SARS-CoV-2 IFN-γ T cell response correlated with S1-SARS-CoV-2-specific serum IgM, IgG, and IgA antibody concentrations. Predominantly, effector memory CD4^+^ T cells of a Th1 phenotype were activated upon exposure to SARS-CoV-2 antigens, which persisted for 4-8 weeks after symptom onset. We detected very low frequencies of SARS-CoV-2-reactive CD8^+^ T cells in these individuals.

**Conclusions:** Our data indicate that an antigen-specific memory CD4^+^ T cell response is induced in children and adults with mild SARS-CoV-2 infection. T cell immunity induced after mild COVID-19 could contribute to protection against re-infection.

## Introduction

Tremendous research efforts have advanced our understanding of immunity to SARS-CoV-2. Most data on the immune response to SARS-CoV-2 was obtained from severe COVID-19 cases [1-4]. However, the vast majority of infected individuals experience mild symptoms that do not require hospitalization [5-8]. The question remains whether individuals, including children, with an asymptomatic or mild SARS-CoV-2 infection, develop immune memory which may protect against subsequent SARS-CoV-2 infections. Persons with mild or asymptomatic infections often develop an antibody response, although not all cases do [8]. It has been shown that SARS-CoV-2-induced antibody levels are waning over time [6, 9-11]. On the other hand, T cell immunity is predicted to persist longer; after SARS-CoV infection in 2003, it was shown that T cell responses can persist for up to 17 years [12]. Some studies investigated the T cell immunity induced after SARS-CoV-2 infection in mild symptomatic adult cases [6, 8, 13-16], showing weaker T cell responses in mild than in moderate or severe COVID-19 cases. CD4^+^ T cell responses against SARS-CoV-2 were more prominent than the CD8^+^ T cell response in adults with mild-to-moderate infection [8, 15, 16], while qualitatively impaired CD4^+^ T cell responses have been reported for critically ill patients [15].

Nevertheless, it remains unclear whether SARS-CoV-2 infection in children, usually showing a mild course, induces substantial T cell immunity. Only a few reports describe the immune responses in children with mild disease or asymptomatic infection, although in these studies T cells specifically reactive to SARS-CoV-2 were not investigated [17-19]. Recently, a study was published investigating SARS-CoV-2 specific T cell responses in children [20]. Induction of a sustainable T cell response is needed to provide immune memory for long-term protection against re-infections by facilitating an efficient and quick response upon re-exposure. Therefore, knowledge on the induction of memory T cell immunity after a mild course of SARS-CoV-2 infection in children and adults is useful for the consideration of the community mitigation measures needed to protect against COVID-19 and limit the spread of the virus.

In the present study, we examined the frequency and the phenotypic/functional characteristics of SARS-CoV-2-reactive T cells in infected children and adults with mild to moderate symptoms. In addition, T cell responses correlated with SARS-CoV-2-specific serum IgM, IgG, and IgA antibody concentrations.

## Methods

### Clinical studies

In this prospective cohort study, described previously [21], households were enrolled in which one adult (index case) tested PCR positive for SARS-CoV-2 between March-May, 2020. Blood samples were collected longitudinally from SARS-CoV-2-infected children and adults of these households. Additionally, blood samples from age-matched unexposed children (n=13) and adults (n=12) were collected from two other cohort studies before the corona pandemic (respectively, 2018-2019 and 2009-2011). The protocol for the SARS-CoV-2-related study, based on the WHO First Few Hundred protocol, was approved by the Medical-Ethical Review Committee of University Medical Center Utrecht (NL13529.041.06). Protocols for the cohort studies with unexposed children (Immfact, NL4679.094.13) and adults (NVI-255, NL29241.000.09) [22] were approved by Medical-Ethical Review Committees of the Netherlands. Written informed consent was received from all participants and/or from parents/guardians of minor participants (<16 years old). All trial-related activities were conducted according to Good Clinical Practice, including the provisions of the Declaration of Helsinki.

### Interferon gamma ELISPOT

Multiscreen filtration ELISPOT plates (Millipore, Merck) were prewetted with 35% ethanol for ≤1 minute and washed with sterile water and PBS. Plates were coated with 5 μg/mL anti-human IFN-γ antibodies (1-D1K, Mabtech) overnight (4°C), then washed with PBS. PBMCs were incubated with heat-inactivated SARS-CoV-2 (MOI-3), or 15-mers overlapping peptides (11 amino acids overlap) covering whole spike protein of SARS-CoV-2 (S-SARS-CoV-2), whole nucleocapsid protein (N-SARS-CoV-2), or S-HCoV-OC43 (0.1 µM/peptide, all JPT), seeded on ELISPOT plates (2.10^5^ cells/well), and incubated for 20 hours, 37°C, 5% CO2 in 100 µl AIM-V (Lonza) with 2% human serum (Sigma). DMSO and PHA (Sigma) were negative and positive controls, respectively. Subsequently, plates were washed and incubated for 1 hour with 1 μg/mL anti-human IFN-γ detection biotinylated-antibody (7-B6-1, Mabtech) in PBS-0.05% casein (Sigma). Plates were washed and incubated with Streptavidin-poly-HRP (Sanquin) in PBS-0.05% casein for 1 hour. After washing, plates were developed with TMB substrate (Mabtech). Spots were analyzed with CTL software. The number of spots from negative controls was subtracted from total spot numbers induced by antigen-specific stimulation; more than 5 spots, after background subtraction, was considered positive. Supernatants and cells from ELISPOT plates were harvested for cytokine release assay and analysis of activation marker expression by T cells, respectively (described in Supplemental Methods).

### Antibody assays

IgM, IgG, and IgA concentrations against SARS-CoV-2 monomeric spike-S1 (40591-V08H; Sino Biological) were determined in serum using a fluorescent bead-based immune assay as published previously [23], with previously determined cutoff values for seroprevalence [10].

### Statistics

Statistical analyses were performed using Prism V7.0 (GraphPad). For unpaired comparisons, Mann-Whitney U test (two groups) or Kruskal-Wallis rank-sum test with Dunn’s posthoc test (≥3 groups) were used. Paired data were compared using the Wilcoxon signed-rank test (two groups) or the Friedman test with Dunn’s multiple comparison test (≥3 groups). Median values for paired comparisons were calculated from subjects with complete data for all time points. Correlation coefficients (r_s_) were determined with Spearman’s rank correlation. Non-parametric tests were used since data were mostly non-normally distributed according to the Shapiro-Wilk test. P values <0.05 were considered significant.

## Results

### Study subjects

SARS-CoV-2 specific T cell responses were assessed from SARS-CoV-2-infected children and adults, and for comparison from unexposed children and adults. Demographic and clinical characteristics are presented in Table 1. Blood samples from 24 children and 27 adults with PCR-confirmed SARS-CoV-2 infection were collected. The median time point of the first sample (T1) was for adults 12.5 days [interquartile range (IQR) 11-14 days] and children 8 days [5.0-16 days] post-symptom onset. Additional blood samples were taken 10-14 days after T1 (referred to as ‘T2’) and only for adults also at 4-6 weeks after T1 (referred to as ‘T3’). No significant differences in major immune cell types were found over time after infection neither in children nor adults, i.e. frequencies of total T cells, B cells, monocytes, or NK cells were comparable between infected groups and healthy age-matched unexposed groups (Supplementary Figure 1).

**Table 1.** Demographic and clinical characteristics of unexposed participants and the cohort of PCR confirmed SARS-CoV-2 infection

| Characteristics | Adults total<br>n = 27 | Adults mild<br>n = 16 | Adults moderate<br>n = 11 | Unexposed adults<br>n = 12 | Children<br>n = 24 | Unexposed children<br>n = 13 |
| --- | --- | --- | --- | --- | --- | --- |
| Median age (range) – years | 44 (18 – 87) | 45 (18 – 87) | 44 (18 – 54) | 38 (20 – 51) | 12 (2 – 16) | 12 (2 – 16) |
| Sex – number (%) |  |  |  |  |  |  |
| Male | 14 (51.9) | 9 (56.3) | 5 (45.5) | 6 (50.0) | 11 (45.8) | 5 (38.5) |
| Female | 13 (48.1) | 7 (43.7) | 6 (54.5) | 6 (50.0) | 13 (54.2) | 8 (61.5) |
| Serology positivity (above cut-off) to Spike-S1 – number (%) |  |  |  |  |  |  |
| IgM | 27 (100.0) | 16 (100.0) | 11 (100.0) | N/A | 20 (83.3) | N/A |
| IgG | 26 (96.3) | 15 (93.7) | 11 (100.0) |  | 19 (79.2) |  |
| IgA | 24 (88.9) | 13 (81.2) | 11 (100.0) |  | 18 (75.0) |  |
| T cell response positivity (≥ 5 spots) to Spike – number (%) |  |  |  |  |  |  |
| IFN-γ ELISPOT | 27 (100.0) | 16 (100.0) | 11 (100.0) | 1 (8.3) | 20 (83.3) | 0 (0.0) |
| Disease severity<sup>A</sup> – number (%) |  |  |  |  |  |  |
| Asymptomatic<sup>B</sup> | 0 (0.0) | 0 (0.0) | 0 (0.0) | N/A | 3 (12.5) | N/A |
| Mild | 16 (59.3) | 16 (100.0) | 0 (0.0) |  | 19 (79.2) |  |
| Moderate/Hospitalized | 11 (40.7) | 0 (0.0) | 11 (100.0) |  | 2 (8.3) |  |
| Sign and symptoms – number (%) |  |  |  |  |  |  |
| Any sign or symptom | 27 (100) | 16 (100.0) | 11 (100.0) | N/A | 21 (87.5) | N/A |
| Fever | 14 (51.9) | 8 (50.0) | 6 (54.5) |  | 6 (25.0) |  |
| Cough | 20 (74.1) | 9 (56.3) | 11 (100.0) |  | 11 (45.8) |  |
| Chills | 15 (55.6) | 8 (50.0) | 7 (63.6) |  | 2 (8.3) |  |
| Sore throat | 14 (51.9) | 9 (56.3) | 5 (45.5) |  | 3 (12.5) |  |
| Runny nose | 18 (66.7) | 11 (68.8) | 7 (63.6) |  | 15 (62.5) |  |
| Phlegm | 12 (44.4) | 7 (43.7) | 5 (45.5) |  | 2 (8.3) |  |
| Headache | 20 (74.1) | 13 (81.2) | 7 (63.6) |  | 7 (29.2) |  |
| Myalgia | 20 (74.1) | 12 (75.0) | 8 (72.7) |  | 1 (4.2) |  |
| Fatigue | 24 (88.9) | 13 (81.2) | 11 (100.0) |  | 6 (25.0) |  |
| Shortness of breath | 9 (33.3) | 0 (0.0) | 9 (81.8) |  | 2 (8.3) |  |
| Loss of taste or smell | 11 (40.7) | 5 (31.2) | 6 (54.5) |  | 3 (12.5) |  |
| Diarrhea | 11 (40.7) | 6 (37.5) | 5 (45.5) |  | 2 (8.3) |  |
| Medical care |  |  |  |  |  |  |
| General practitioner | 5 (18.5) | 2 (12.5) | 3 (27.3) | N/A | 2 (8.3) | N/A |
| Hospitalization | 7 (25.9) | 0 (0.0) | 7 (63.6) |  | 0 (0.0) |  |
| Comorbidities – number (%) |  |  |  |  |  |  |
| Total | 14 (51.9) | 7 (43.7) | 7 (63.6) | 0 (0.0) | 7 (29.2) | 6 (46.2) |
| Cardiovascular disease | 2 (7.4) | 1 (6.3) | 1 (9.1) | 0 (0.0) | 0 (0.0) | 0 (0.0) |
| Lung disease | 4 (14.8) | 0 (0.0) | 4 (36.4) | 0 (0.0) | 4 (16.7) | 3 (23.1) |
| Proteinuria | 1 (3.7) | 0 (0.0) | 1 (9.1) | 0 (0.0) | 0 (0.0) | 0 (0.0) |
| Lupus | 1 (3.7) | 1 (6.3) | 0 (0.0) | 0 (0.0) | 0 (0.0) | 0 (0.0) |
| Osteoporosis | 1 (3.7) | 1 (6.3) | 0 (0.0) | 0 (0.0) | 0 (0.0) | 0 (0.0) |
| Allergies | 5 (18.5) | 4 (25.0) | 1 (9.1) | 0 (0.0) | 6 (25.0) | 5 (38.5) |

<sup>A</sup>Disease severity was classified as asymptomatic (absence of symptoms), mild (at least one symptom but absence of shortness of breath), moderate (presence of shortness of breath with or without other symptoms, including hospitalized cases). None of the hospitalized patients were admitted to an intensive care unit, thus not considered severe cases. <sup>B</sup>Three children were remained asymptomatic during the study but tested PCR positive. n, number of subjects in specific group; Spike-S1, SARS-CoV-2 Spike S1 protein; Spike, overlapping peptides of SARS-CoV-2 spike protein; N/A, not applicable

### SARS-CoV-2 specific IFN-γ^+^ T cell response

IFN-γ^+^-producing cells were detected by ELISPOT upon stimulation with overlapping peptides covering spike protein (S) of SARS-CoV-2 (S-SARS-CoV-2) in 83% (20/24) of infected children and in 100% (27/27) of infected adults, whereas IFN γ^+^ responses were found in 0% (0/6) and 8.3% (1/12) of the unexposed children and adults, respectively. Frequencies of SARS-CoV-2-specific IFN-γ^+^ T cells were lower in infected children than in infected adults (Figure 1A-C); the median spot forming units (SFU)/2.10^5^ PBMCs for infected children vs infected adults was 18 vs 62 (P=0.0021) at T1 upon stimulation with S-SARS-CoV-2 (Figure 1A). At T1, a 2-to 6-fold higher frequency of SARS-CoV-2-specific IFN-γ^+^ T cells was found, depending on the antigenic stimulus used, in adults with moderate symptoms compared to mild symptomatic adults (upon S-SARS-CoV-2-specific stimulation, 105 vs 45 SFU/2.10^5^ PBMCs at T1 (P=0.045) (Figure 1F-G)). At later time points, higher frequencies of IFN-γ-producing T cells in moderately ill patients compared to mild symptomatic adults were only observed after stimulation with overlapping peptides covering nucleocapsid protein (N) of SARS-CoV-2 (N-SARS-CoV-2) at T3 or with inactivated whole SARS-CoV-2 at T2 and T3. The higher frequencies of SARS-CoV-2-specific IFN-γ^+^ T cells observed in infected adults compared to children were mainly caused by the higher responses found in adults with moderate symptoms. Nevertheless, the IFN-γ response of mild symptomatic adults also tended to be slightly higher than infected children (Figure 1E-F). The pre-existing T cell response against S-HCoV-OC43 of infected children was very low, although S-HCoV-OC43-specific T cell frequency was slightly higher at T2 compared to the age-matched unexposed control group (9.0 vs 1.0 SFU/2.10^5^ PBMCs; P=0.037) (Figure 1D, left panel). In infected adults, no difference in numbers of S-HCoV-OC43-specific T cells was found between SARS-CoV-2-infected and unexposed adults (Figure 1D, right panel). In both SARS-CoV-2-infected children and adults, the frequency of IFN-γ^+^ T cells was 4 to 60-fold lower after stimulation with S-HCoV-OC43 (Figure 1D) compared to stimulation with any of the SARS-CoV-2 antigens (Figure 1A-C). No significant difference in frequency of S-HCoV-OC43-reactive T cells was observed between unexposed adults and unexposed children or between mild and moderate COVID-19 cases.

**Figure 1.**
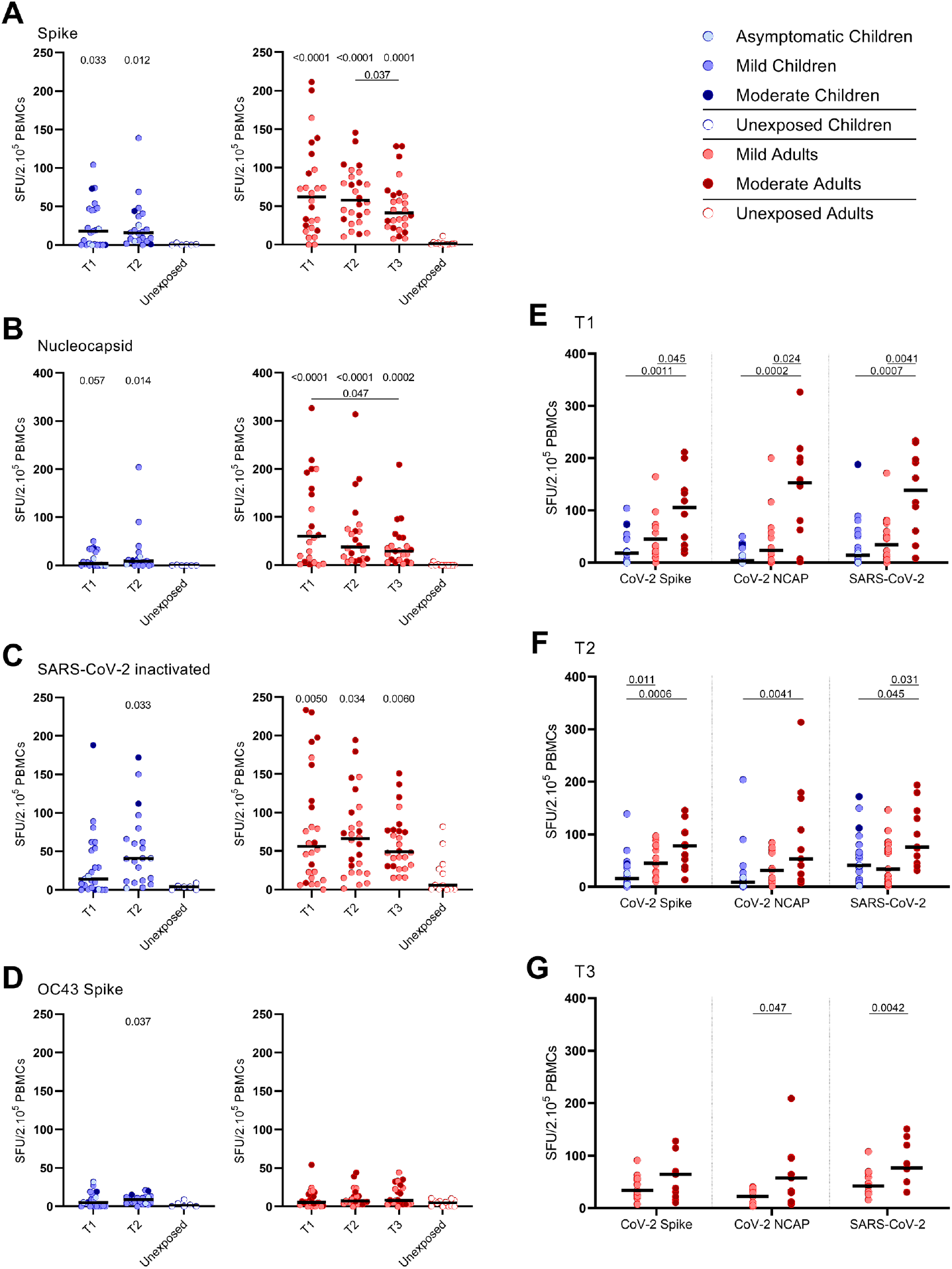
SARS-CoV-2-specific IFN-γ^+^ T cell response in infected children and infected adults (mild and moderate cases) versus unexposed healthy controls over time after infection. Dot plots summarizing the frequencies of IFN-γ-producing cells responding to SARS-CoV-2 and HCoV-OC43 antigens for (A-D) children (left panel), adults (right panel), over time after infection, and compared to unexposed adults/children (ELISPOT assay). Frequencies of IFN-γ-producing cells responding to (A) set of overlapping of peptides of SARS-CoV-2 spike protein, (B) set of overlapping peptides of SARS-CoV-2 nucleocapsid protein, (C) inactivated SARS-CoV-2, and (D) set of overlapping peptides of HCoV-OC43 spike protein. (E) Comparison of IFN-γ-producing cells derived from infected children, mild symptomatic SARS-CoV-2-infected adults versus adult COVID-19 patients with moderate symptoms at T1, (F) at T2, and (G) mild symptomatic SARS-CoV-2-infected adults versus adult COVID-19 patients with moderate symptoms at T3. Each dot represents one subject. Bars indicate the median of spot-forming units per 200,000 PBMCs. SFU, spot-forming unit. (A-D) P values related to comparisons with the unexposed controls are listed at the top of the graph, above the corresponding group for comparison. For unpaired comparisons, Mann-Whitney U test (two-group comparisons) (mild adults versus moderate adults) or Kruskal-Wallis rank-sum test with Dunn’s posthoc test for multiple comparisons were used (children versus mild adults versus moderate adults; unexposed versus infected children or adults at T1 versus T2 versus T3). Differences between paired data were compared using the Wilcoxon signed-rank test (for comparison of two paired groups) (infected children at T1 versus T2) or the Friedman test with Dunn’s multiple comparison tests (infected adults at T1 versus T2 versus T3). Statistically significant comparisons are indicated, with P values < 0.05 considered significant. T1, first timepoint of sampling for adults median 12.5 days and children median 8 days post-symptom onset; T2, 10-14 days after T1; T3, 4-6 weeks after T1.

### Activation of effector memory CD4^+^ T cells upon SARS-CoV-2-specific stimulation

Especially the CD25 (IL-2Rα) and CD137 (4-1BB) expression on T cells of infected subjects increased significantly upon the various SARS-CoV-2 antigenic stimulations compared to mock stimulation (Supplementary Figure 2). Primarily CD4^+^ T cells and not CD8^+^ T cells expressed CD25/CD137 activation markers upon SARS-CoV-2 antigenic stimulation, in infected subjects (Figure 2A-G, Supplementary Figure 3). Compared to the unexposed age-matched groups, higher frequencies of activated CD4^+^ T cells were observed in both infected children and infected adults after stimulation with any of the three SARS-CoV-2 antigen preparations. Frequencies of SARS-CoV-2-specific activated CD4^+^ T cells were significantly lower in infected children than in adults (upon S-SARS-CoV-2-specific stimulation, 0.04% CD25^+^/CD137^+^ T cells of total CD4^+^ T cells for infected children vs 0.21% for infected adults at T1; P=0.0035) (Figure 2A-G). Frequencies of SARS-CoV-2-specific activated CD4^+^ T cells, irrespective of used SARS-CoV-2 antigen, were higher in the adults with moderate COVID-19 illness compared to adults with mild symptoms at T1. This difference was not observed at later time points (Figure 2A-G). The SARS-CoV-2-specific activated CD4^+^ T cells were mainly effector memory (CD45RO^+^/CCR7^-^) (T_EM_) (Figure 2H).

**Figure 2.**
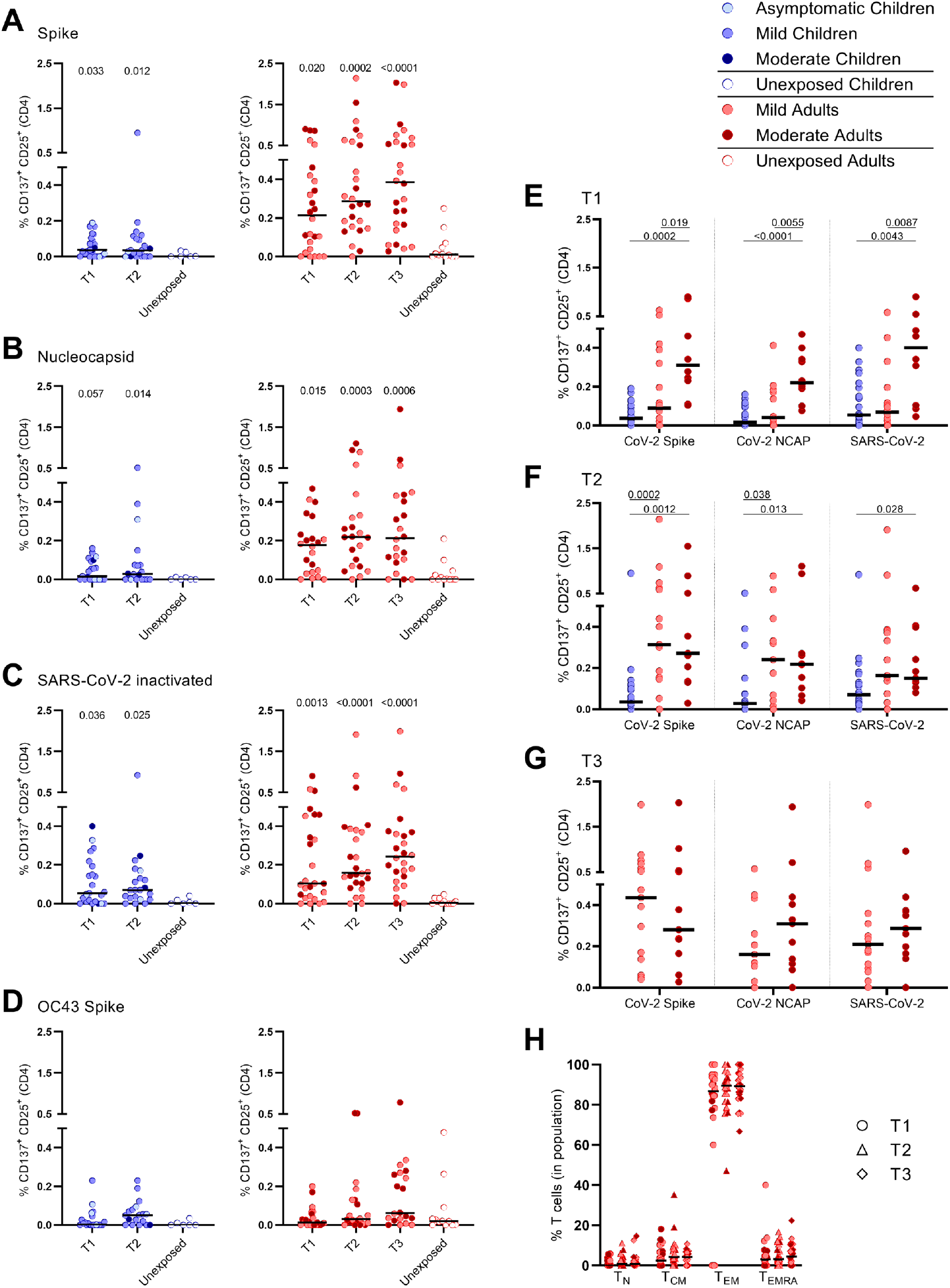
Frequencies of activated CD4^+^ T cells of infected children and infected adults (mild and moderate cases) versus unexposed healthy controls over time after infection. Dot plots summarizing the percentages of CD25^+^/CD137^+^ activated CD4^+^ T cells responding to SARS-CoV-2 and HCoV-OC43 antigens for (A-D) children (left panel), adults (right panel), over time after infection, and compared to unexposed adults/children. Percentages of CD25^+^/CD137^+^ activated CD4^+^ T cells responding to (A) set of overlapping peptides of SARS-CoV-2 spike protein, (B) set of overlapping peptides of SARS-CoV-2 nucleocapsid, (C) inactivated SARS-CoV-2, and (D) set of overlapping peptides of HCoV-OC43 spike protein. (E) Comparison of IFN-γ-producing cells derived from infected children, mild symptomatic SARS-CoV-2-infected adults versus adult COVID-19 patients with moderate symptoms at T1, (F) at T2, and (G) mild symptomatic SARS-CoV-2-infected adults versus adult COVID-19 patients with moderate symptoms at T3. (H) Immunophenotyping at the single-cell level showed the different memory subsets within the SARS-CoV-2-specific activated CD4^+^ T cells from infected adults. Each dot represents one subject. Bars indicate the median percentage of total CD4^+^ T cells. (A-D) P values related to comparisons with the unexposed controls are listed at the top of the graph, above the corresponding group for comparison. For unpaired comparisons, Mann-Whitney U test (two-group comparisons) (mild adults versus moderate adults) or Kruskal-Wallis rank-sum test with Dunn’s posthoc test for multiple comparisons were used (children versus mild adults versus moderate adults; unexposed versus infected children or adults at T1 versus T2 versus T3). Differences between paired data were compared using the Wilcoxon signed-rank test (for comparison of two paired groups) (infected children at T1 versus T2) or the Friedman test with Dunn’s multiple comparison tests (infected adults at T1 versus T2 versus T3). Statistically significant comparisons are indicated, with P values < 0.05 considered significant.T1, first timepoint of sampling for adults median 12.5 days and children median 8 days post-symptom onset; T2, 10-14 days after T1; T3, 4-6 weeks after T1.

### Correlations between SARS-CoV-2-specific IFN-γ^+^ T cell responses and CD4^+^ T cell response

In children, moderate correlations were observed between IFN-γ^+^ T cell frequency and activated (CD137^+^/CD25^+^)CD4^+^ T cells after stimulation with both S-SARS-CoV-2 (r_s_=0.45; P=0.036) and N-SARS-CoV-2 (r_s_=0.64; P=0.004) at T2, and at T1 only after stimulation with inactivated SARS-CoV-2 (r_s_=0.51; P=0.012) (Figure 3A). In adults, IFN-γ^+^ T cell frequency and activated (CD137^+^/CD25^+^)CD4^+^ T cells correlated after stimulation with both S-SARS-CoV-2 and N-SARS-CoV-2 at all three time points after infection (r_s_ ranging between 0.43-0.66) and after stimulation with inactivated SARS-CoV-2 at T1 and T2 (respectively, r_s_=0.66 and r_s_=0.53) (Figure 3B).

**Figure 3.**
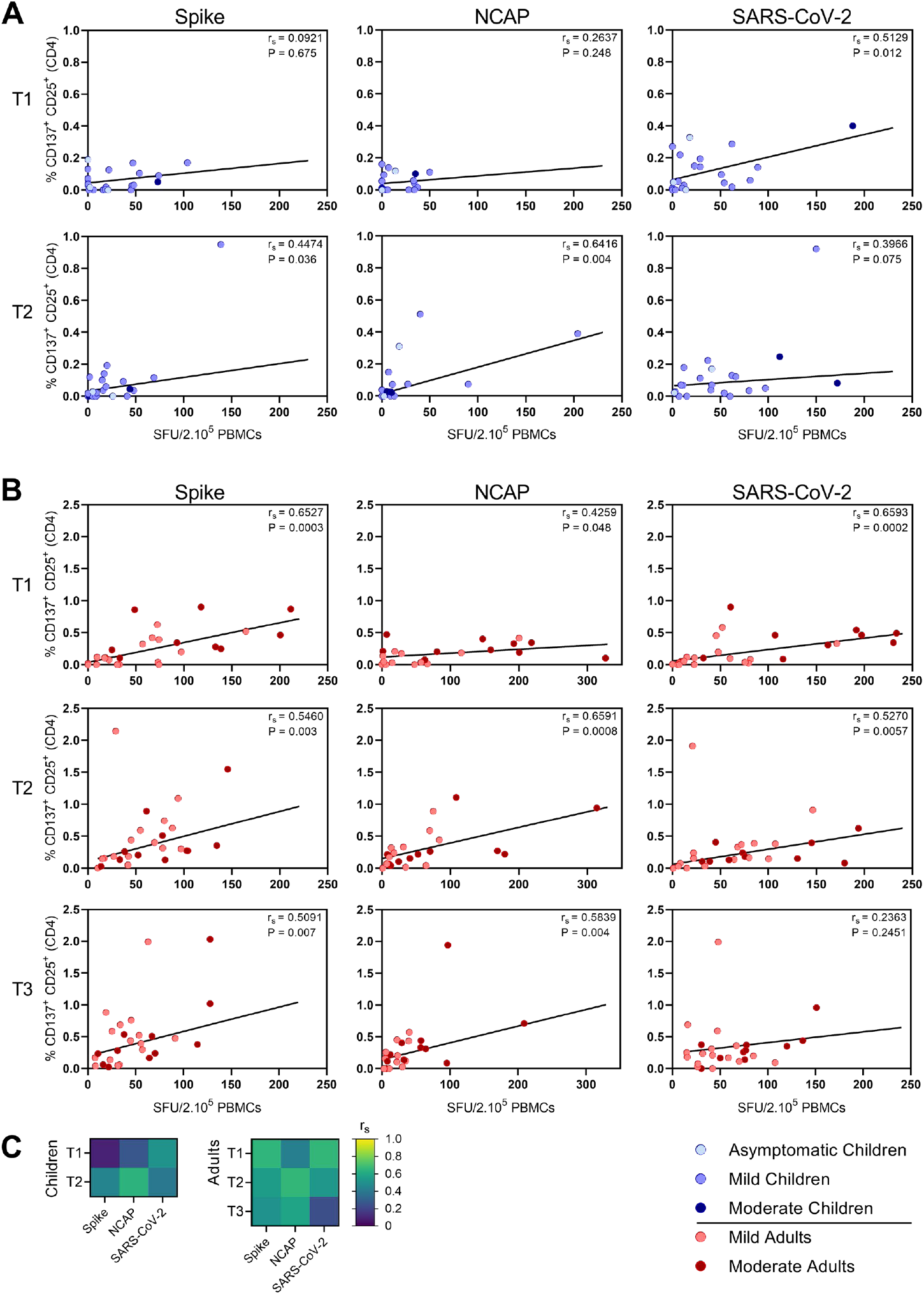
Correlation between IFN-γ^+^ T cell frequency and activated CD4^+^ T cells. Spearman correlation between frequency of IFN-γ^+^ responder cells and percentages of CD25^+^/CD137^+^ activated CD4^+^ T cells of (A) children and (B) adults responding to a set of overlapping peptides of SARS-CoV-2 spike protein (left panel), or a set of overlapping peptides of SARS-CoV-2 nucleocapsid protein (middle panel) or inactivated SARS-CoV-2 (right panel) at different time points after infection. (C) Heatmaps summarizing the pairwise correlations. Each dot represents one subject. Correlation coefficients (r_s_) were determined with Spearman’s rank correlation. P values < 0.05 were considered significant. T1, first timepoint of sampling for adults median 12.5 days and children median 8 days post-symptom onset; T2, 10-14 days after T1; T3, 4-6 weeks after T1.

### SARS-CoV-2-specific release of cytokines

Upon stimulation with S-SARS-CoV-2, PBMCs from SARS-CoV-2-infected children secreted more IL-2 than unexposed children, albeit at very low amounts (3.7 vs 0.1 pg/ml; P=0.030), as well as IL-10 (1.5 vs 0.1 pg/ml; P = 0.019). Similar trends were observed in SARS-CoV-2-infected adults compared to unexposed adults, for both IL-2 (18.0 vs 0.1 pg/ml; P=0.0003) and IL-10 (2.6 vs 0.1 pg/ml; P=0.0087) secretion. S-SARS-CoV-2-specific IL-2 secretion was higher in infected adults compared to infected children (P=0.015). However, IL-2 secretion was only significantly higher in adults with moderate COVID-19 (51.8 vs 3.7 pg/ml; P=0.0026) and not in adults with mild symptoms compared to infected children (Figure 4). Other cytokines were only secreted at low levels and not significantly.

**Figure 4.**
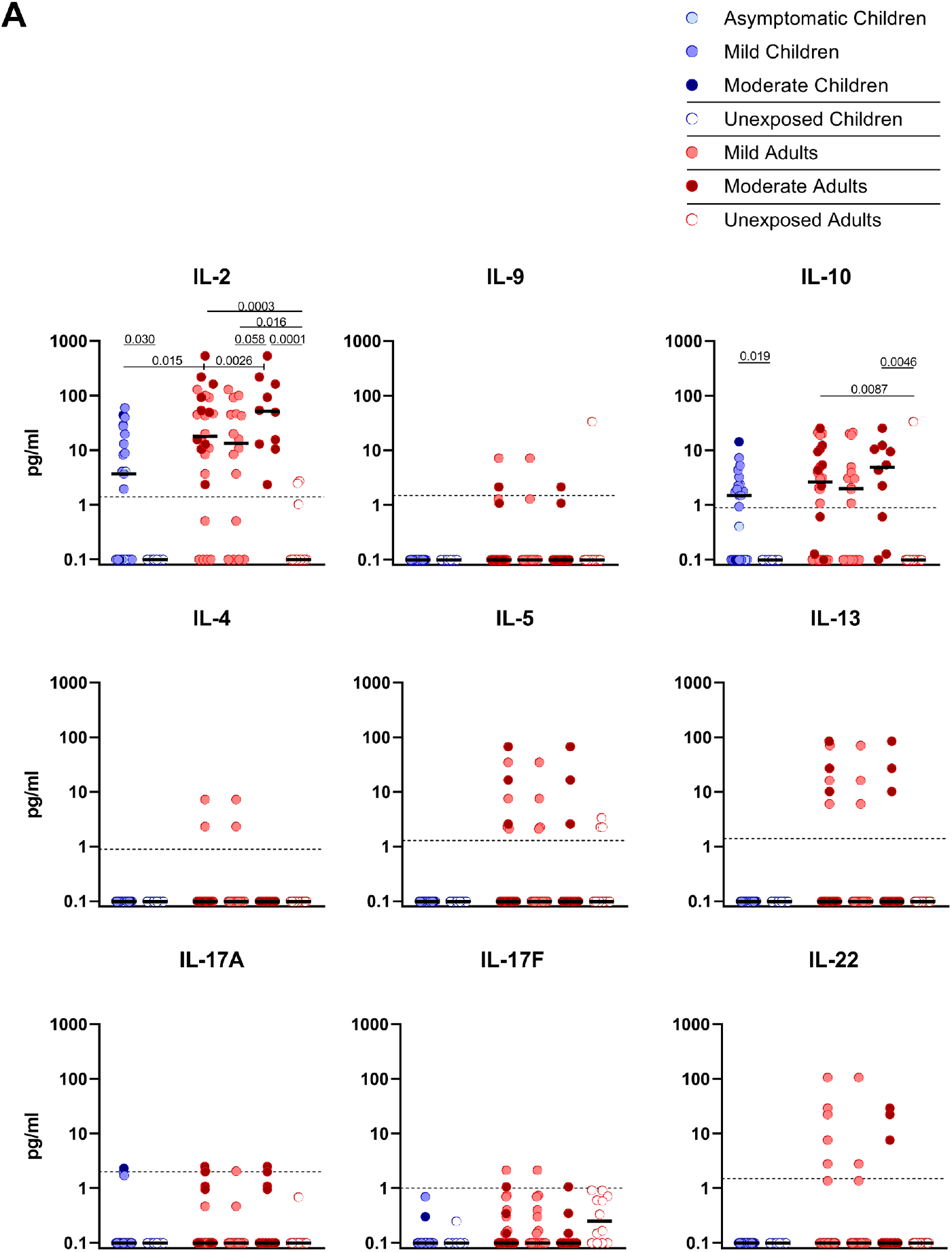
Cytokine release in infected children and mild versus moderate symptomatic adults over time after infection. Cell-free culture supernatants were harvested from IFN-γ ELISPOT plates and the release of the following cytokines was measured in T1 samples: IL-2, IL-4, IL-5, IL-9, IL-10, IL-13, IL-17A, IL-17F, IL-22. Dot plots show the concentration of cytokines (pg/ml) after stimulation of PBMCs with a set of overlapping peptides of SARS-CoV-2 spike protein. Infected children and unexposed children versus infected adults with mild or moderate disease and unexposed adults are depicted. Minimum detection threshold (MDT) concentrations for each cytokine, as calculated by the manufacture and mentioned in Supplemental Methods, are indicated with horizontal dotted lines. Each dot represents one subject. Bars indicate the median cytokine concentration (pg/ml). For two-group comparisons (infected children versus unexposed children), Mann-Whitney U test was used. Kruskal-Wallis rank-sum test with Dunn’s posthoc test for multiple comparisons was used (all adults vs mild adults versus moderate adults versus unexposed adults; all adults vs mild adults versus moderate adults versus infected children). P-values ≤ 0.05 are presented.

### Correlations between SARS-CoV-2-specific IFN-γ^+^ T cell frequency and antibody response

Serum antibody concentrations against the SARS-CoV-2 Spike S1 protein (S1-SARS-CoV-2) above the previously established cutoff level [10] at any of the two sampling time points were found in 83.3% (IgM), 79.2% (IgG), and 75.0% (IgA) of the infected children. From the four children without detectable S-SARS-CoV-2-specific T cell responses, two did have S1-SARS-CoV-2-specific IgM, IgG, and IgA antibodies; the other two did not. In adults, 100%, 96.3%, and 88.9% were seropositive for respectively, IgM, IgG, and IgA antibodies to S1-SARS-CoV-2 at any of the sampling time points. Interestingly, in children good correlations were observed between S-SARS-CoV-2 IFN-γ^+^ T cell frequency and S1-SARS-CoV-2-specific serum IgM, IgG and IgA concentrations, though this was only observed at T1 (for IgM, R_s_=0.73 (P<0.0001); IgG, r_s_=0.74 (P<0.0001); IgA, r_s_=0.66 (P=0.0005)) (Figure 5A). In adults, frequency of S-SARS-CoV-2-specific T cells was also correlated with S1-SARS-CoV-2-specific serum IgM concentrations at T2 and T3 (respectively, r_s_=0.42 (P=0.03) and r_s_=0.49 (P=0.009)), with S1-SARS-CoV-2-specific serum IgG concentrations at T2 and T3 (for both time points, r_s_=0.47 (P=0.01)), and with S1-SARS-CoV-2-specific serum IgA concentrations at T3 (r_s_=0.44 (P=0.02)) (Figure 5B).

**Figure 5.**
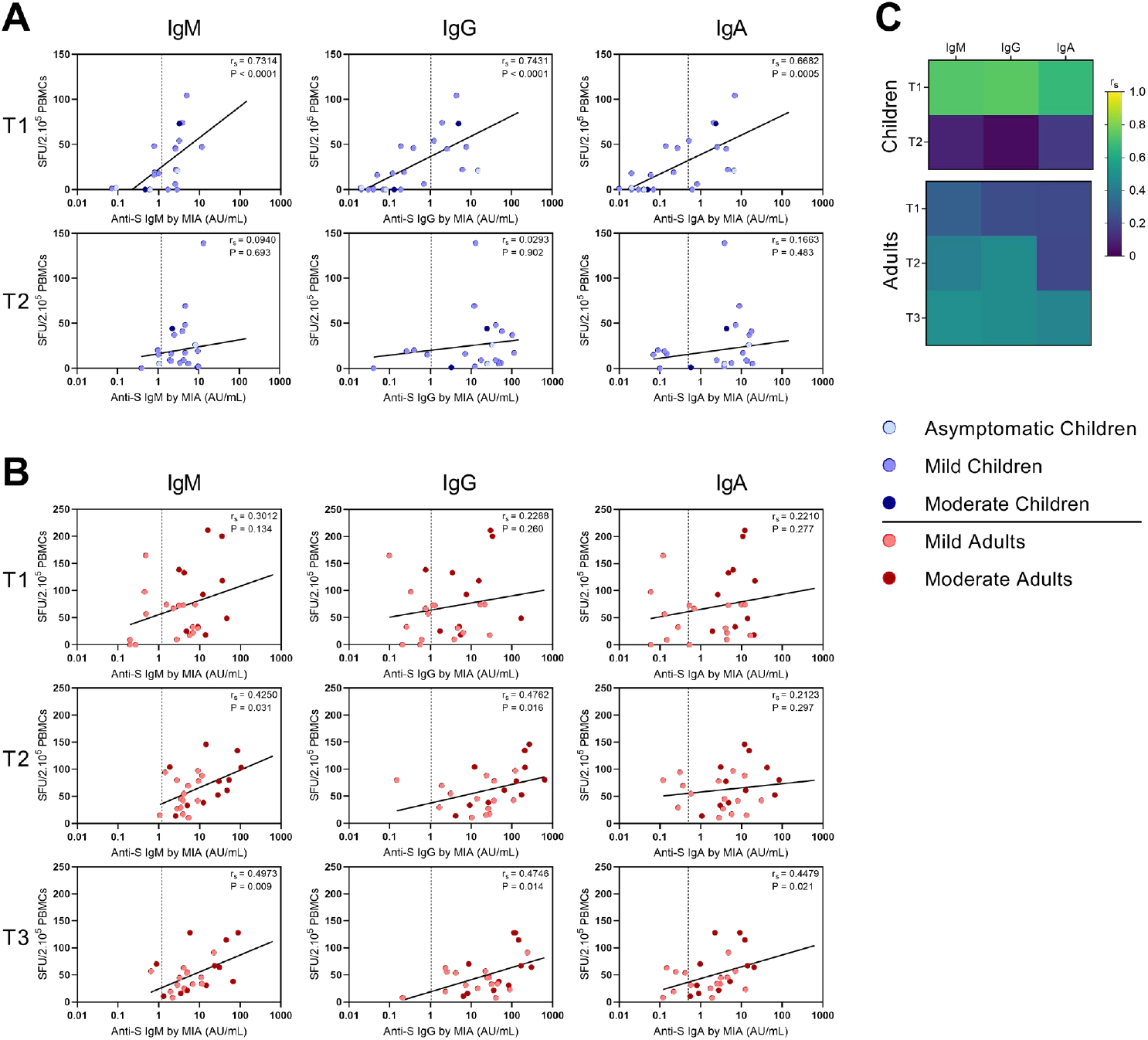
Correlation between IFN-γ^+^ T cell frequency and antibody concentrations to S-SARS-CoV-2. Spearman correlation between frequency of IFN-γ^+^ responder cells and concentrations of Spike-SARS-CoV-2 IgM, IgG, or IgA antibodies at different time points after infection in (A) children and (B) adults. (C) Heatmaps summarizing the pairwise correlations. Cut-off values for seroprevalence are 1.20, 1.04, and 0.50 AU/ml for Spike-SARS-CoV-2-specific IgM, IgG, and IgA, respectively, and are indicated with vertical dotted lines. Each dot represents one subject. Correlation coefficients (r_s_) were determined with Spearman’s rank correlation. P values < 0.05 were considered significant. T1, first timepoint of sampling for adults median 12.5 days and children median 8 days post-symptom onset; T2, 10-14 days after T1; T3, 4-6 weeks after T1.

## Discussion

Most infections with SARS-CoV-2, especially among children, have a mild course. But, do children, despite experiencing mild infection, develop memory T cell immunity? Here, we describe the kinetics, function, and phenotype of SARS-CoV-2-specific T cells of infected children in comparison with adults experiencing mild to moderate COVID-19 symptoms. Only limited studies have been reported investigating the immune responses in children with mild/asymptomatic SARS-CoV-2 infection [17-20]. The strength of our study is that we evaluated the recall T cell response upon SARS-CoV-2 specific stimulation, and compared it to unexposed children and adults.

We found higher frequencies of SARS-CoV-2-specific IFN-γ^+^ T cells in all infected groups compared to the unexposed control groups after any of the three antigenic stimulations that included heat-inactivated SARS-CoV-2, and overlapping peptides of SARS-CoV-2 spike protein (S-SARS-CoV-2), and nucleocapsid protein (N-SARS-CoV-2). In general, frequencies of IFN-γ^+^-T cells reactive against SARS-CoV-2 antigens were lower in infected children, who generally had mild/asymptomatic infection, compared to infected adults. The lower SARS-CoV-2-specific T cell response observed in children suggests that other compartments of the immune system, such as the innate immune response, contribute to faster clearing of the infection, as demonstrated by others [24-26]. The higher T cell responses in infected adults could largely be explained by higher T cell responses found in moderate cases, although adults with mild complaints also tended to have slightly higher responses than infected children. This is in agreement with findings from other studies showing higher frequencies of SARS-CoV-2-specific T cells in severe patients compared to mildly symptomatic patients [8, 15]. In contrast, critically ill patients have been reported to exhibit qualitatively impaired S-SARS-CoV-2-specific CD4^+^ T cell responses, indicating that a good CD4^+^ T cell response may protect against serious disease [15]. It has been demonstrated that SARS-CoV-2-specific T cell responses can be retained >8 months following infection regardless of disease severity [7, 16, 27, 28]. In line with these results, we also do find memory responses, which could be indicative of sustained T cell immunity after mild SARS-CoV-2 infection. In agreement with our IFN-γ ELISPOT data, significantly lower frequencies of SARS-CoV-2-specific activated (CD25^+^/CD137^+^/)CD4^+^ T cells were observed in infected children compared to infected adults. Recently, Cohen et al. (2021) also described that acute and memory CD4^+^ T cells in children were significantly lower than in adults, while polyfunctional cytokine production by T cells was comparable [20]. In accordance with Cohen et al. [20], we found that the SARS-CoV-2 activated T cells mainly belonged to the CD4^+^ effector memory subset (T_EM_: CD45RO^+^/CCR7^-^). Data on the SARS-CoV-2-specific IFN-γ^+^ T cell response and CD4^+^ T cell activation correlated, suggesting that IFN-γ was produced by antigen-specific CD4^+^ T cells. Apart from IFN-γ, SARS-CoV-2-specific T cells produced IL-2, suggesting a Th1 phenotype of the CD4^+^ T_EM_.

In contrast, very low frequencies of activated (CD25^+^/CD137^+^) CD8^+^ T cells were detected after SARS-CoV-2-specific stimulation. An explanation for this may be that CD8^+^ T cells have migrated to the local sites of infection to attack virus-infected cells or might be a result of the antigenic stimulation used in our study. Smaller peptides (9 to 10-mers instead of 15-mers) or live SARS-CoV-2 may be more suitable to measure CD8^+^ T cell responses. Sekine et al. also observed proportionately larger SARS-CoV-2-specific CD4^+^ T cell responses than CD8^+^ T cell responses to different sets of overlapping peptides in the convalescent phase of both mild and severe COVID-19 cases, although in that study also IFN-γ^+^ CD8^+^ T cell responses were detected [8].

Pre-existing cross-reactive T cell immunity generated by common cold human coronaviruses (HCoV) has been suggested to affect clinical outcomes of SARS-CoV-2 infection. T cell lines from unexposed healthy donors specific for S-HCoV-229E and -OC43 were cross-reactive to S-SARS-CoV-2 [29]. Based on these findings, the authors suggested that children may have higher HCoV prevalence due to more frequent social contacts, explaining their lower risk for severe COVID-19 [29]. In the present study, we did, however, not find a significant difference in the low frequencies of S-HCoV-OC43-reactive IFN-γ^+^ T cells between unexposed children and unexposed adults. In another study with mild COVID-19 adult patients, also low T cell frequencies recognizing S-HCoV-229E/S-HCoV-OC43 peptide pools were found [15]. It cannot be excluded that pre-existing cross-reactive immunity to other conserved parts of SARS-CoV-2 played a role, or that pre-existing immunity to other HCoV played a role. We, however, found no evidence that pre-existing S-HCoV-OC43-reactive T cells boosted upon SARS-CoV-2 infection could explain the mild course of infection.

We found positive correlations between SARS-CoV-2-specific IFN-γ^+^ T cell frequency and serum-IgG, -IgM, and -IgA antibody concentrations to S1-SARS-CoV-2 in both children and adults. Although, it should be taken into account that the antibody concentrations of considerable numbers of children were below the threshold for seropositivity [10]. Other studies have also shown positive correlations between S-SARS-CoV-2-specific IgG antibodies and T cell responses [6, 30, 31].

Limited data is available on the immune response to SARS-CoV-2 in children. Here, we show significantly higher frequencies of SARS-CoV-2-specific T cells in infected children compared to unexposed age-matched controls. Frequencies of SARS-CoV-2 reactive T cells were lower in infected children whom almost all had mild/asymptomatic infection compared to infected adults with mild to moderate COVID-19. Predominantly CD4^+^ T cells, and not CD8^+^ T cells, were activated upon stimulation with SARS-CoV-2 antigens. SARS-CoV-2-specific T cell responses were more prominent in infected adults with moderate symptoms than with mild infection. Importantly, our data indicate that an effector memory T cell response is developed after experiencing mild SARS-CoV-2 infection. It is tempting to speculate that such responses may contribute at least partially to protection against re-infection and might limit the spread of the virus. A follow-up study is planned to evaluate the long-term T cell response.

## Data Availability

All relevant data are included in the article or in the Supplementary Materials. Additional data is available upon request respecting the privacy of the participants and other restrictions related the data.

## Acknowledgments

We are grateful to the members of the Dutch FFX-COVID-19 research group of the Centre for Infectious Disease Control, National Institute for Public Health and the Environment, the Netherlands, that includes Arianne van Gageldonk-Lafeber, Wim van der Hoek, Susan van den Hof, Chantal Reusken and Inge Roof for their contribution to the design and the conduct of the SARS-CoV-2-related clinical study. We thank the Public Health Service Utrecht for assistance in the recruitment of households. We thank Anneke Westerhof, Anne-Marie van den Brandt, Anoek Backx, Bas van der Veer, Elma Smeets -Roelofs, Elsa Porter, Elske Bijvank, Fion Brouwer, Francoise van Heiningen, Gabriel Goderski, Gert-Jan Godeke, Harry van Dijken, Helma Lith, Hinke ten Hulscher, Ilse Akkerman, Ilse Schinkel, Jeroen Hoeboer, Johan Reimerink, Jolanda Kool, Josien Lanfermeijer, Joyce Greeber, Kim Freriks, Kina Helm, Lidian Izeboud, Lisa Wijsman, Liza Tymchenko, Martijn Vos, Margriet Bisschoff, Marieke Hoogerwerf, Marit de Lange, Marit Middeldorp, Marjan Bogaard, Marjan Kuijer, Nening Nanlohy, Olga de Bruin, Rob van Binnendijk, Rogier Bodewes, Ronald Jacobi, Ruben Wiegmans, Sakinie Misiedjan, Saskia de Goede, Sharon van den Brink, Sophie van Tol, Teun Guichelaar, Titia Kortbeek, and Yolanda van Weert for support during the stages of this study, household visits, processing samples and laboratory analyses.

