## Supplemental Data for "Children and adults with mild COVID-19 symptoms develop memory T cell immunity to SARS-CoV-2"

### Supplemental Methods

#### Peripheral blood mononuclear cells and serum isolation

PBMCs were isolated from heparinized blood by centrifugation on a Ficoll-Hypaque gradient (Pharmacia Biotech) and cryopreserved at -135°C until use. Serum was separated from the blood by clotting and centrifugation and stored at −80°C until analysis.

#### Ex vivo immune profiling

Defrosted PBMCs from SARS-CoV-2 infected children (n=24) and adults (n=27), as well as from unexposed healthy children (n=13) and adults (n=12), were used for deep immune profiling by multicolor flow cytometry. (BD FACSymphony™). Cells were stained for anti-human CD3 (OKT3), CD14 (HCD14), CD28 (CD28.2), CD56 (5.1H11), CD57 (HNK-1), CD95 (DX2) and CCR7 (G043H7) (all Biolegend) and CD4 (RPA-T4), CD8 (RPA-T8), CD19 (SJ25C1), CD27 (L128), CD45RO (UCHL1) (all BD Bioscience) and eFluor 780 fixable viability stain (65-0865-14, ThermoFisher). For this purpose, major lymphocyte populations were discriminated by analyzing the CD3 expression to identify T cells , and identifying $CD4^+$, $CD8^+$, $CD4^+/CD8^+$ and $CD4^-/CD8^-$ T cells within the $CD3^+$ T cells, analyzing CD19 expression to detect B cells, CD56 expression for NK cells and CD14 expression to identify cells of the myelomonocyte lineage. Memory $CD4^+$ and $CD8^+$ T cell subsets were further discriminated based on CD45RO and CCR7 staining (true naïve T cells ($T_N$), $CD45RO^-$, $CD27^+$, $CCR7^+$, $CD95^-$; central memory T cells ($T_{CM}$), CD45RO$^+$, CD27$^+$; effector memory T cells ($T_{EM}$), CD45RO$^+$, CD27$^-$; terminally differentiated effector memory T cells re-expressing CD45RA ($T_{EMRA}$), CD45RO$^-$, CD27$^-$, CD28$^-$, CD57$^+$). Flowcytometry data analysis was performed using FlowJo software, version 10 (TreeStar).

**Generation of heat-inactivated virus stocks of SARS-CoV-2**

SARS-CoV-2 isolate, hCoV-19/Netherlands/Zuid_Holland_0133R/2020, was obtained from a Dutch patient. Virus was grown on VERO-E6 cells in DMEM medium (Gibco; Thermo Fisher Scientific) supplemented with 1x penicillin-streptomycin-glutamine (Gibco) and 2% FBS for approximately 48 hours under BSL-3 conditions. At >90% cytopathic effect (CPE), the suspension was collected and spun down (4000 × g, 10 min) to remove cell debris. Virus stocks were aliquoted and stored at −80°C until use. The 50% tissue culture infective dose ($TCID_{50}$), $7.63.10^7$ $TCID_{50}$/ml, was determined by the Reed and Muench method. Heat-inactivation was performed by incubating the virus at 60 °C for 2 hours, after which the inactivated virus stocks were stored at −80°C until use.

**Immunophenotyping and expression of activation markers after in vitro stimulation**

Activated T cells were determined by harvesting cells from the IFN-ɣ ELISPOT plates and subsequently flow cytometric analysis using activation markers. Cells were stained for anti-human CD3 (SK7), CD4 (SK3), CD8 (RPA-T8), CD45RO (UCHL1), CD25 (2A3), CD56 (NCAM16.2), CD69 (FN50), OX40 (L106)and fixable viability stain 780 (BD Bioscience) and CCR7 (G043H7) (Biolegend). After fixation and permeabilization, using FoxP3/Transcription Factor Staining Buffer Set (eBioscience, Thermo Fisher Scientific, Waltham, Mass) cells were stained intracellularly for anti-human, CD137 (4-1BB) and CD154 (BD Bioscience). Data were acquired on a FACS Symphony analyzer (BD) and analyzed using FlowJo (V10, Tree Star, Ashland, Ore).

**Cytokine release assay**

Cell-free culture supernatants were harvested from the IFN-ɣ ELISPOT plates, and analyzed using a multiplex bead-based assay quantitating levels of IL-2, IL-4, IL-5, IL-6, IL-9, IL-10, TNF, IL-13, IL-17A, IL-17F, IL-22 (LEGENDplex; BioLegend) according to the manufacturers' instructions and using

FACSCanto (BD). For analysis, the online cloud-based program, The LEGENDplex™ Data Analysis Software Suite, was used. The minimum detection threshold (MDT) for each cytokine as calculated by the manufacture was: 1.4 pg/ml for IL-2, 0.9 pg/ml for IL-4, 1.3 pg/ml for IL-5, 1.1 pg/ml for IL-6, 1.5 pg/ml for IL-9, 0.9 pg/ml for IL-10, 0.9 pg/ml for TNF, 1.4 pg/ml for IL-13, 2.0 pg/ml for IL-17A, 1.0 pg/ml for IL-17F, and 1.5 pg/ml for IL-22. Background signal for IL-6 and TNF from unstimulated controls was as high as stimulated samples, therefore these cytokines were excluded from analysis.

### Supplementary Figure 1

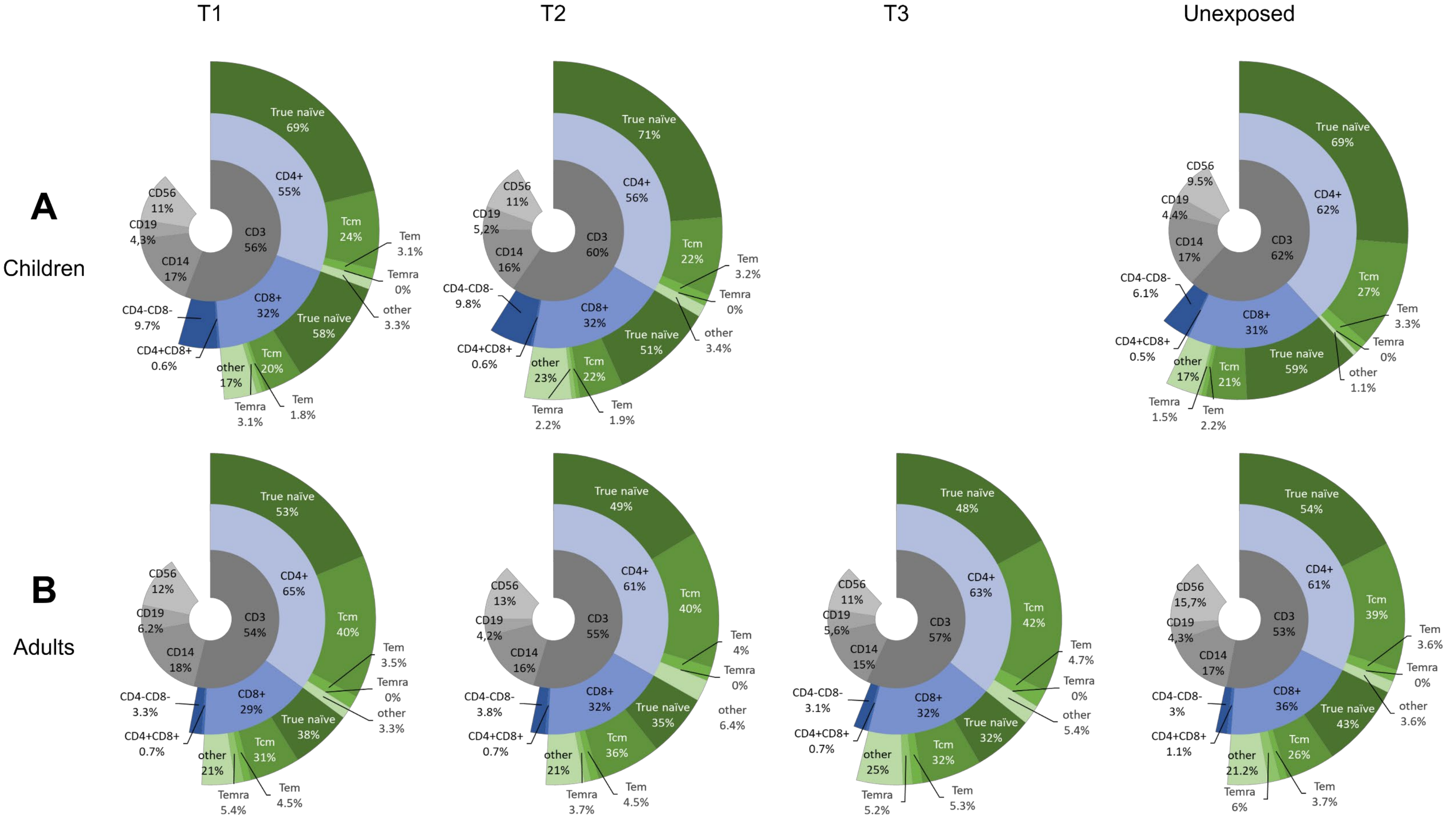

**Supplementary Figure 1. *Ex vivo* immune profiling of PBMCs of adults and children over time after infection**

Sunburst plots showing the results of immunophenotyping of PBMCs performed with multicolor flow cytometry by staining for: (inner circle, lymphocyte gate) B cells ($CD19^+$, $CD3^-$); NK cells ($CD56^+$, $CD3^-$, $CD19^-$); $CD14^+$ monocytes ($CD14^+$, $CD3^-$, $CD19^-$, $CD56^-$), T cells ($CD3^+$); (middle circle, $CD3^+$ T cells) $CD4^+$ T cells ($CD4^+$, $CD8^-$); $CD8^+$ T cells ($CD8^+$, $CD4^-$); $CD4^+$/$CD8^+$ T cells ($CD3^+$, $CD4^+$); $CD4^-$/$CD8^-$ T cells ($CD4^-$, $CD8^-$); (outer circle) $CD4^+$ and $CD8^+$ memory T cell subsets are identified based on CD45RO, CCR7, CD27, CD28 and CD95 staining (true naive, $CD45RO^-$, $CD27^+$, $CCR7^+$, $CD95^-$; TCM, $CD45RO^+$, $CD27^+$; $T_{EM}$, $CD45RO^+$, $CD27^-$; $T_{EMRA}$, $CD45RO^-$, $CD27^-$, $CD28^-$, $CD57^+$), as indicated. Children in time after SARS-CoV-2 infection and unexposed children (A); Adults in time after SARS-CoV-2 infection and unexposed healthy adults (B). Data are presented as median percentages of presented cell types.

Presented median percentages of T cell subsets in the sunburst plots may slightly differ from the median percentages described in the text as for comparison between the various time points, as described in the text, only paired data were considered.

T1, first timepoint of sampling for adults median 12.5 days and children median 8 days post-symptom onset; T2, 10-14 days after T1; T3, 4-6 weeks after T1.

Supplementary Figure 2

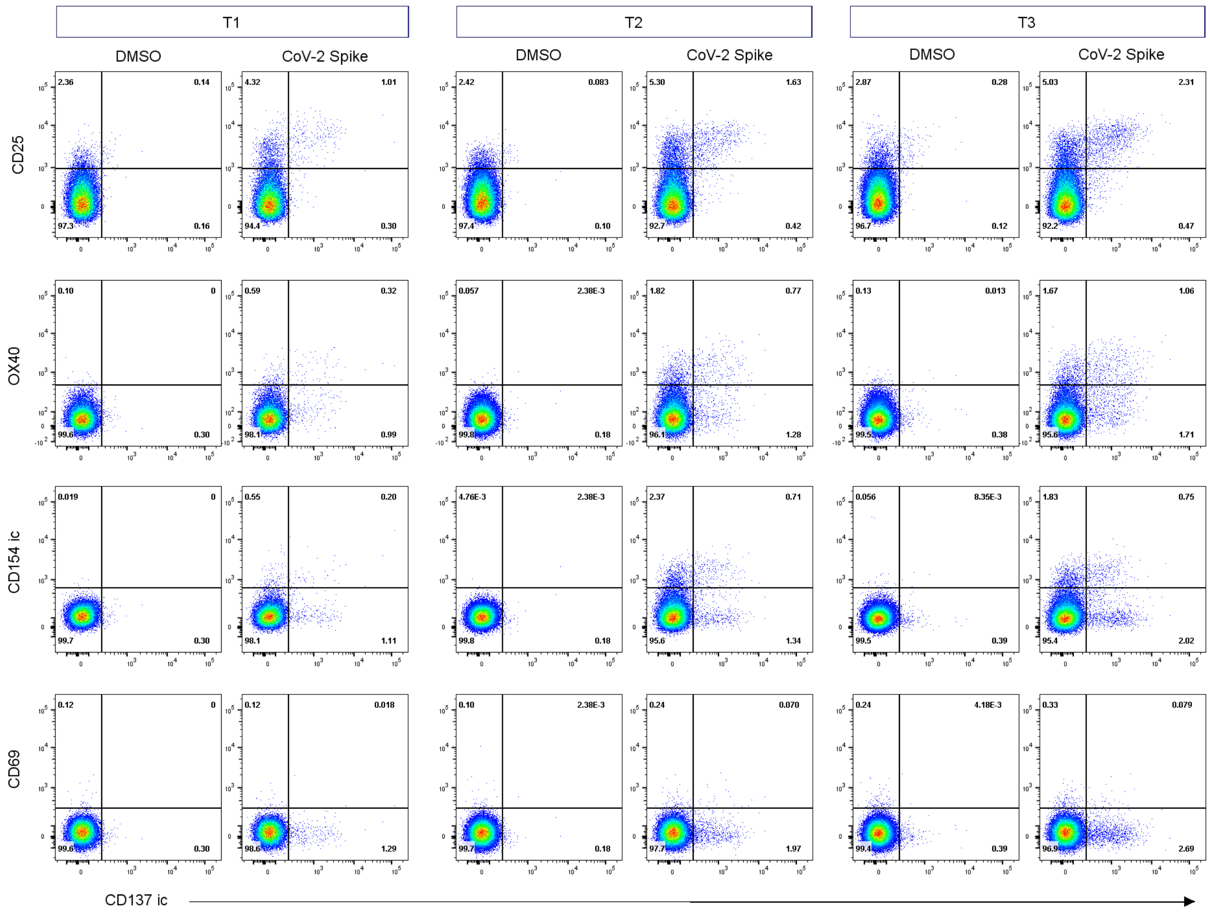


**Supplementary Figure 2. Expression of different activation markers on spike-SARS-CoV-2-stimulated CD4$^{+}$ T cells**

Representative flow cytometry plots of expression of various activation markers on CD4$^{+}$ T cells 24 hours after incubation of PBMCs from infected adults with overlapping peptides of SARS-CoV-2 spike protein versus DMSO (negative control) in time after infection, T1 (left panel), T2 (middle panel), and T3 (right panel). The following activation markers were used: CD25, OX40, CD154 (intracellular staining), CD69, and CD137 (intracellular staining).

T1, first timepoint of sampling for adults median 12.5 days and children median 8 days post-symptom onset; T2, 10-14 days after T1; T3, 4-6 weeks after T1

### Supplementary Figure 3

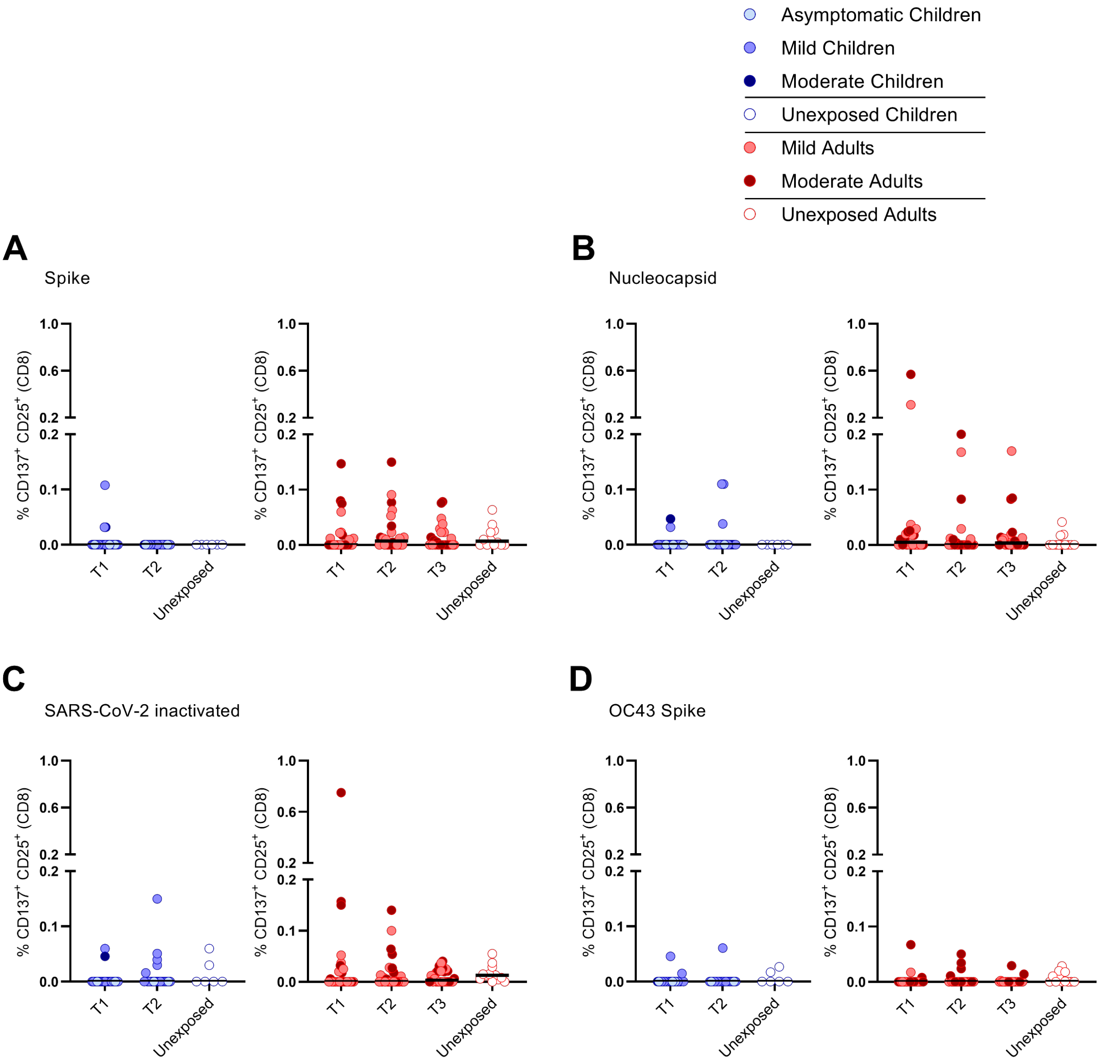


**Supplementary Figure 3. Percentages of activated CD8⁺ T cells in adults and children over time after infection versus unexposed healthy controls**

Dot plots summarizing the frequencies of $CD25^+/CD137^+$ activated $CD8^+$ T cells responding to SARS-CoV-2 and HCoV-OC43 antigens for children versus unexposed controls (left panel), adults versus unexposed controls (right panel) in time after infection. Frequencies of $CD25^+/CD137^+$ activated $CD8^+$ T cells responding to (A) inactivated SARS-CoV-2, (B) overlapping peptides of SARS-CoV-2 spike protein, (C) overlapping peptides of SARS-CoV-2 nucleocapsid protein, and (D) overlapping peptides of HCoV-OC43 spike protein.

Each dot represents one subject. Bars indicate median of % $CD25^+$/$CD137^+$ $CD8^+$ T cells. For unpaired comparisons, Kruskal-Wallis rank-sum test with Dunn's posthoc test for multiple comparisons was used (infected children at T1 or T2 versus unexposed children, infected adults at T1 or T2 versus unexposed adults). Differences between paired data were compared using the Wilcoxon signed-rank test (for comparison of two paired groups) (infected children at T1 versus T2) or the Friedman test with Dunn's multiple comparison tests (infected adults at T1 versus T2 versus T3). Statistically significant comparisons are indicated, with P values < 0.05 considered significant.

T1, first timepoint of sampling for adults median 12.5 days and children median 8 days post-symptom onset; T2, 10-14 days after T1; T3, 4-6 weeks after T1.
